# Estimating the elevated transmissibility of the B.1.1.7 strain over previously circulating strains in England using GISAID sequence frequencies

**DOI:** 10.1101/2021.03.17.21253775

**Authors:** Chayada Piantham, Natalie M. Linton, Hiroshi Nishiura, Kimihito Ito

## Abstract

The B.1.1.7 strain, also referred to as Alpha variant, is a variant strain of the severe acute respiratory syndrome coronavirus 2 (SARS-CoV-2). The Alpha variant is considered to possess higher transmissibility compared to the strains previously circulating in England. This paper proposes a new method to estimate the selective advantage of a mutant strain over another strain using the time course of strain frequencies and the distribution of the serial interval of infections. This method allows the instantaneous reproduction numbers of infections to vary over calendar time. The proposed method also assumes that the selective advantage of a mutant strain over previously circulating strains is constant. Applying the method to SARS-CoV-2 sequence data from England, the instantaneous reproduction number of the B.1.1.7 strain was estimated to be 26.6–45.9% higher than previously circulating strains in England. This result indicates that control measures should be strengthened by 26.6–45.9% when the B.1.1.7 strain is newly introduced to a country where viruses with similar transmissibility to the preexisting strain in England are predominant.

## Introduction

Severe acute respiratory syndrome coronavirus 2 (SARS-CoV-2), the causative agent of COVID-19, has rapidly evolved since its introduction to the human population in 2019. In December 2020, Public Health England detected a new cluster of SARS-CoV-2 viruses phylogenetically distinct from the other strains circulating in the United Kingdom (Chand et al. 2020). These viruses were assigned the lineage name B.1.1.7 following the Phylogenetic Assignment of Named Global Outbreak Lineages (PANGO) nomenclature (Rambaut et al. 2020b). The World Health Organization (WHO) designated the lineage as a variant of concern (VOC) in December 2020, and it is now known as VOC Alpha (World Health Organization 2020).

It was retrospectively determined that the B.1.1.7 strain was first detected in England in September 2020, and the number of infections with this strain increased in October and November in 2020 (Chand et al. 2020). By February 2021, the B.1.1.7 strain accounted for 95% of SARS-CoV-2 circulation in England (Davies et al. 2021).

Several studies have compared the transmissibility of the B.1.1.7 strain to that of previously circulating strains. Davies et al. estimated that the reproduction number *R* (the average number of secondary infections generated by a given primary infection) of the B.1.1.7 strain is 43–90% (95% credible interval [CrI]: 38–130%) higher than preexisting strains (Davies et al. 2021). However, other models found different ranges of estimates in their multiplicative increase in *R*. Grabowski et al. estimated a 83–118% increase with a confidence interval of 71–140% compared to previously circulating strains in England (Grabowski et al. 2021). Volz et al. estimated a 50–100% increase in *R* using data from England (Volz et al. 2021), while Washington et al. estimated a 35–45% increase using data from the United States using Volz’s method (Washington et al. 2021). As well, Chen et al. estimated a 49–65% increase using data from Switzerland (Chen et al. 2021). Strong control measures including movement restrictions and ban on meeting and event were taken to respond to the introduction of a strain with high transmissibility. Thus, the *R* may change over time during the course of an epidemic during which new SARS-CoV-2 variant strains emerge.

In this paper, we propose a method to estimate the selective advantage of a mutant strain over previously circulating strains. Based on Fraser’s method to estimate the instantaneous reproduction number using a renewal equation (Fraser 2007), our method allows the reproduction number to vary over calendar time. Our approach is also based on the Maynard Smith’s model of allele frequencies in adaptive evolution, which assumes that the selective advantage of a mutant strain over previously circulating strains is constant over time (Maynard Smith and Haigh 1974). Applying the developed method to the sequence data in England using the serial interval distribution of COVID-19 estimated by Nishiura et al. (Nishiura et al. 2020), we estimated the change in the instantaneous reproduction number of B.1.1.7 strains compare to that of strains previously circulating in England.

## Materials and Methods

### Sequence data

Nucleotide sequences of SARS-CoV-2 viruses were obtained from the GISAID EpiCoV database (Shu and McCauley 2017) on March 1, 2021. Nucleotide sequences of viruses detected in England were selected and aligned to the reference amino acid sequence of the spike protein of SARS-CoV-2 (YP_009724390) using DIAMOND (Buchfink et al. 2015). The aligned nucleotide sequences were translated into amino acid sequences, then were aligned with the reference amino acid sequence using MAFFT (Katoh et al. 2002). Amino acid sequences having either an ambiguous amino acid or more than ten gaps were excluded from the rest of analyses. Table 1 shows the amino acids on the spike protein used to characterize the B.1.1.7 strain, as retrieved from the PANGO database (Rambaut et al. 2020b).

**Table 1.** Amino acids on the spike protein which are used to define B.1.1.7 strains.

| Position on S protein | Amino acid |
| --- | --- |
| 69 | Deletion |
| 70 | Deletion |
| 144 | Deletion |
| 501 | Tyrosine (Y) |
| 570 | Aspartic acid (D) |
| 681 | Histidine (H) |
| 716 | Isoleucine (I) |
| 982 | Alanine (A) |
| 1118 | Histidine (H) |

We divided amino acid sequences into three groups based on the amino acids shown in Table 1. The first group consists of sequences having all the B.1.1.7-defining amino acids in Table 1. We call a virus in this group a “B.1.1.7 strain”. The second group contains sequences that have none of the B.1.1.7-defining substitutions. We call a virus in this group a “non-B.1.1.7 strain”. The third group is a set of sequences that have at least one but incomplete set of the B.1.1.7-defining amino acids. We call a strain in the third group a “B.1.1.7-like strain”. Table 2 shows the number of sequences categorized into each group. Figure 1 shows the daily numbers of GISAID sequences of B.1.1.7 strains, non-B.1.1.7 strains and B.1.1.7-like strains detected in England from September 1, 2020 to February 19, 2021. We used the number of B.1.1.7 strains and non-B.1.1.7 strains for the rest of the analyses. B.1.1.7-like strains were excluded from the analyses as it is unclear whether they have the same transmissibility as B.1.1.7 strains. These numbers are provided in Supplementary Table 1.

**Table 2.** Number of GISAID sequences in England from September 1, 2020 to February 19, 2021.

| B.1.1.7 strains | Non-B.1.1.7 | B.1.1.7-like | Total |
| --- | --- | --- | --- |
| 71,692 | 65,840 | 2,227 | 139,759 |

**Figure 1.**
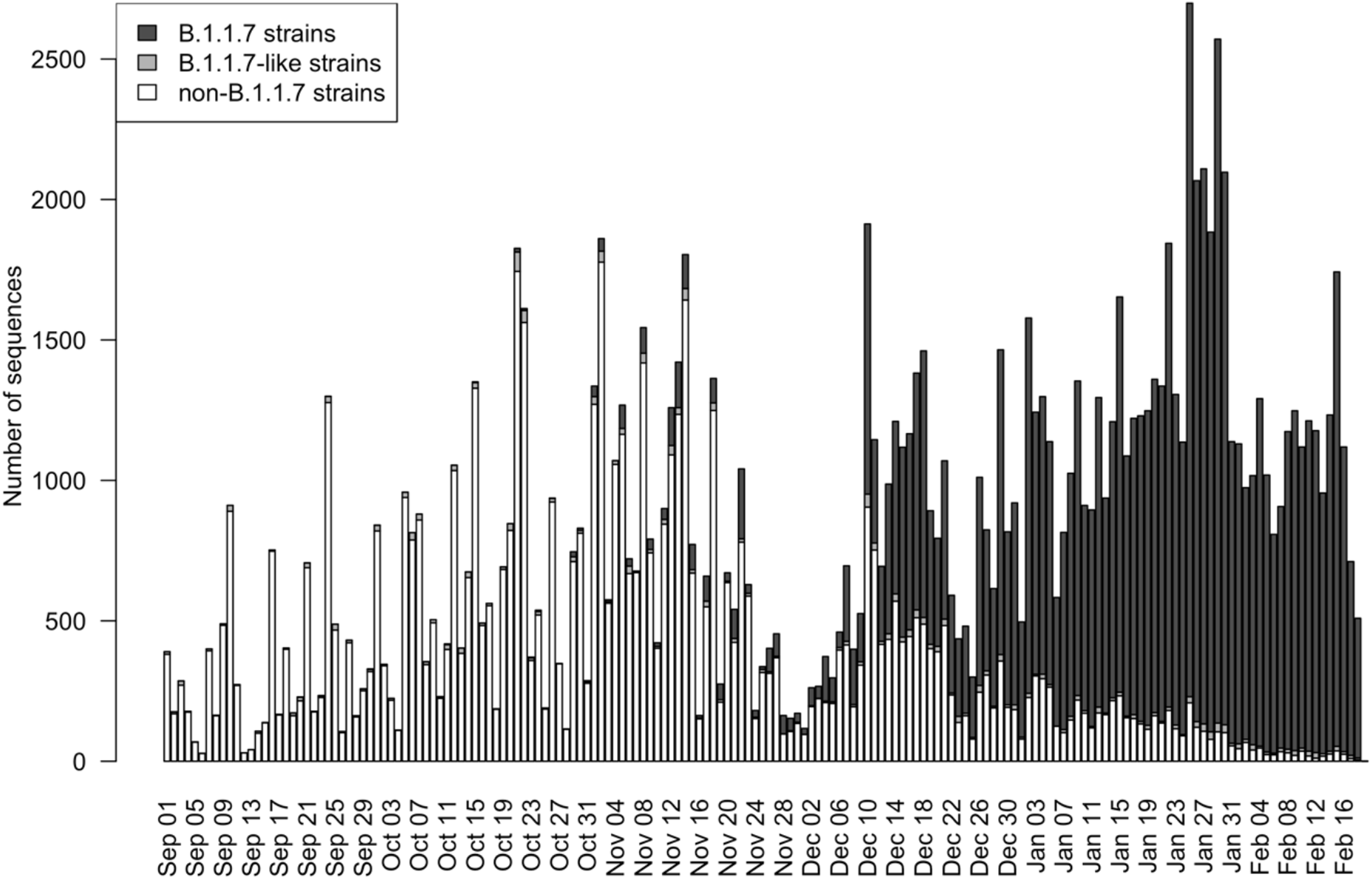
Numbers of nucleotide sequences of B.1.1.7 strains, B.1.1.7-like strains, and non-B.1.1.7 strains in England from September 1, 2020 to February 19, 2021, based on sequences retrieved from the GISAID database on March 1, 2020.

### Serial interval distribution

The serial interval is the time from illness onset in a primary case to illness onset in a secondary case (Nishiura et al. 2020). The method we propose in this paper uses discrete distributions of serial intervals. The discretized probability mass function of the serial interval for *i* ≥ 0 days is given by

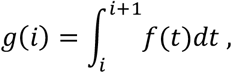

where *f*(*t*) is the probability density function of a lognormal distribution with a log mean of *μ* and log standard deviation of *σ*. Values of *μ* and *σ* were estimated by maximizing the likelihood of parameters, using datasets of illness onset among infector–infectee pairs labeled with “certain” and “probable” in the dataset published by Nishiura et al. (Nishiura et al. 2020).

### Model of Advantageous Selection

Let us suppose that we have a large population of viruses consisting of strains of two genotypes *A* and *a*, of which frequency in the viral population at a calendar date *t* are *q*_*A*_(*t*) and *q*_*a*_(*t*), respectively. Suppose also that genotype *A* is a mutant of *a* that emerged at time *t*_0_.

We assume that a virus of genotype *A* generates 1 + *s* times as many secondary transmissions as those of genotype *a*. Then, *s* can be considered as the coefficient of selective advantage in adaptive evolution. As described in Maynard Smith and Haigh (1974), the frequency of viruses of allele *A* after *n* transmissions, *q*_*n*_, satisfies the following equation:

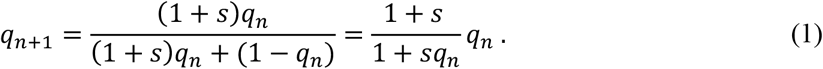

Maynard Smith’s formulation of allele frequency can be extended using the concept of instantaneous reproduction numbers of infectious diseases. The instantaneous reproduction number is defined as the average number of people someone infected at time *t* could expect to infect given that conditions remain unchanged (Fraser 2007). Let *I*(*t*) be the number of infections by viruses of either genotype *A* or *a* at calendar time *t* and *g*(*i*) be the probability mass function of serial intervals, defined in the previous subsection. Suppose that instantaneous reproduction numbers of genotypes *A* and *a* at calendar time *t* are *R*_*A*_(*t*) and *R*_*a*_(*t*), respectively. Assuming that the distribution of generation time of a disease can be approximated by the serial interval distribution, the following equations give the discrete version of Fraser’s instantaneous reproduction numbers of infections by genotype *A* and *a* at time *t*.

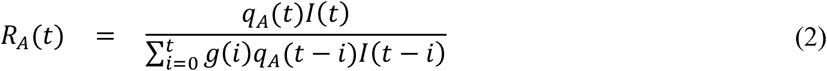

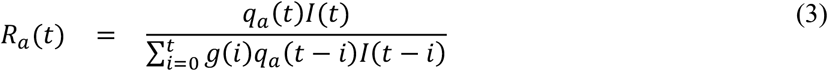

Suppose that *g*(*i*) is small enough to be neglected for *i* < 1 and *i* > *l*, then *g*(*i*) can be truncated and the above formula can be treated as follows.

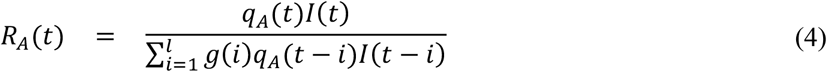

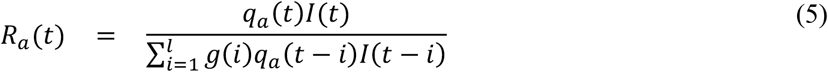

Since a virus of genotype *A* generates 1 + *s* times as many secondary transmissions as those of genotype *a*, the following equation holds

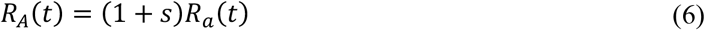

for each calendar time *t* ≥ *t*_0_. Next we assume that for all infections at calendar time *t*, the difference in the number of infections at the time when previous generations became infected can be regarded as considerably small, i.e.

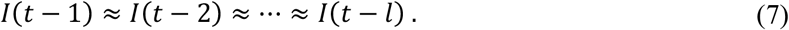

The frequency of genotype *A* in the viral population at calendar time *t, q*_*A*_(*t*), can be modeled as follows:

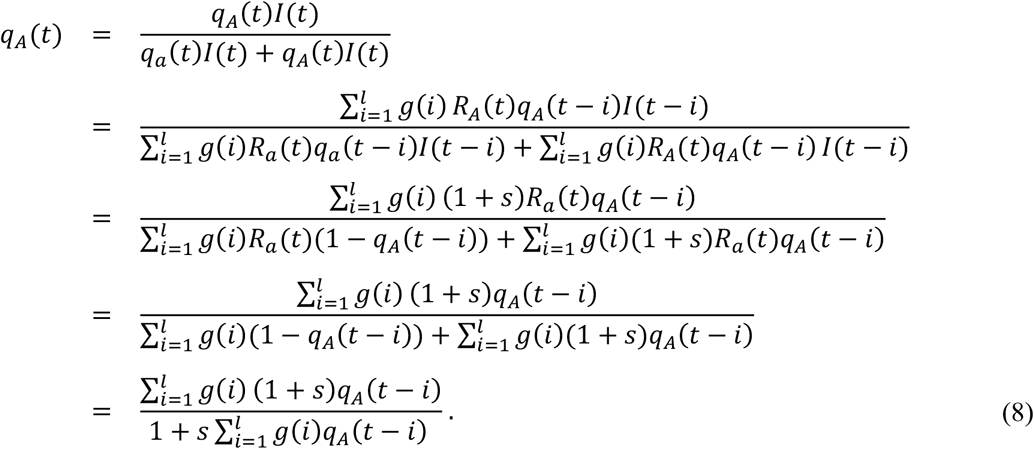

### Likelihood Function

Let *n*(*t*) be the number of sequences of either genotype *A* or *a* observed at calendar date *t*. Let *d*_1_, …, *d*_*k*_ be calendar dates such that *n*(*d*_*i*_) > 0 for 1 ≤ *j* ≤ *k*. Suppose that we have *n*_*A*_(*d* _*j*_) sequences of genotype *A* at calendar date *d*_*j*_. Since genotype *A* emerged at time *t*_0_, *q*_*A*_(*d*_*j*_) =0 *and q*_*a*_(*d*_*j*_) = 1 for *d*_*j*_ < *t*_0_. If the is frequency of genotype *A* is *q*_*A*_(*t*_0_), then the following equation gives the likelihood function of *s, t*_0_, and *q*_*A*_(*t*_0_) for observing *n*_*A*_(*d*_*j*_) sequences of genotype *A* at calendar date *d*_*j*_:

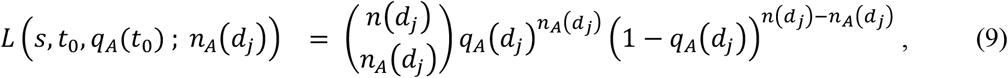

for 1 ≤ *j* ≤ *k*. The likelihood function of *s, t*_0_, and *q*_0_ for observing *n*_*A*_(*d*_1_), …, *n*_*A*_(*d*_*k*_) sequences of genotype *A* at calendar dates *d*_1_, …, *d*_*k*_ is given by the following formula.

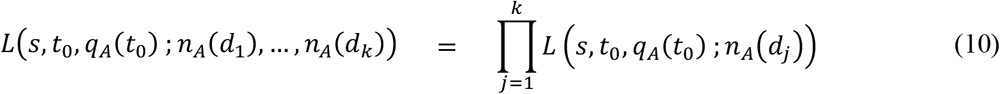

### Parameter estimation from sequence data

The B.1.1.7 strain was first detected in England on September 20, 2020. We assume that *t*_0_ is this day or someday before this day. Parameters *s, t*_0_, and *q*(*t*_0_) were estimated by maximizing the likelihood of observations on September 1, 2020 and later on. B.1.1.7 strains. Viruses having complete subset of B.1.1.7-defining substitutions on its spike protein were considered as genotype *A*. The non-B.1.1.7 strains, viruses having none of B.1.1.7-defining substitutions were considered genotype *a*. The B.1.1.7-like strains, viruses having an incomplete set of B.1.1.7 substitutions on the spike protein, were excluded from analysis. We truncated the distribution of serial intervals so that *g*(*i*) =0 if *i* < 1 or *i* > 20 and normalized *g*(*i*) to ensure that 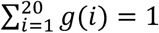. Parameters of *s, t*_0_, and *q*(*t*_0_) were estimated by maximizing the likelihood defined in Equation (10). The 95% confidence intervals of parameters were estimated by profile likelihood (Pawitan 2013). Optimization of the likelihood function was performed using the nloptr package in R (Johnson 2020; Rowan 1990). Effects of the log mean and standard deviation of serial interval distribution on the estimate of selective advantage *s* were evaluated using the bootstrap-based random samples of *μ* and *σ* that were taken from the boundary of 95% confidence area on the likelihood surface for the serial interval distribution.

## Results

The selective advantage of B.1.1.7 strains over non-B.1.1.7 strains, *s*, was estimated at 0.344 (95% confidence interval [CI] 0.343 to 0.346) (Table 2). These estimates were obtained by assuming that the serial intervals follow the lognormal distribution with a log mean of 1.38 and log standard deviation of 0.56 based on the empirical serial interval data. The date when a B.1.1.7 virus have emerged in England (*t*_0_) was estimated to be September 20, 2020 (95% CI: September 17–20). The initial frequency of B.1.1.7 among non-B.1.1.7 and B.1.1.7 strains at the emergence in England, *q*_0_, was estimated to be 0.00556 with its 95% confidence intervals from 0.00534 to 0.00581.

**Table 2.** Maximum likelihood estimations of parameters.

| Parameter | Estimated value* | 95% CI |
| --- | --- | --- |
| $s$ | 0.344 | [0.343, 0.346] |
| $t_0$ | September 20, 2020 | [September 17, 2020, September 20, 2020] |
| $q_0$ | 0.00556 | [0.00534, 0.00581] |

\* Estimates and 95% CIs were obtained by assuming that the serial intervals followed the lognormal distribution with a log mean of 1.38 and log standard deviation of 0.56.

Figure 2 shows the temporal change in the frequency of B.1.1.7 strains among all strains except B.1.1.7-like strains detected in England from September 1, 2020 to February 19, 2021. White circles indicate daily frequencies of B.1.1.7 strains among all strains except B.1.1.7-like strains. Solid line indicates the time course of frequency of B.1.1.7 strains calculated using parameters estimated from the data. Dashed lines indicate its lower and upper bounds of its 95% CI.

**Figure 2.**
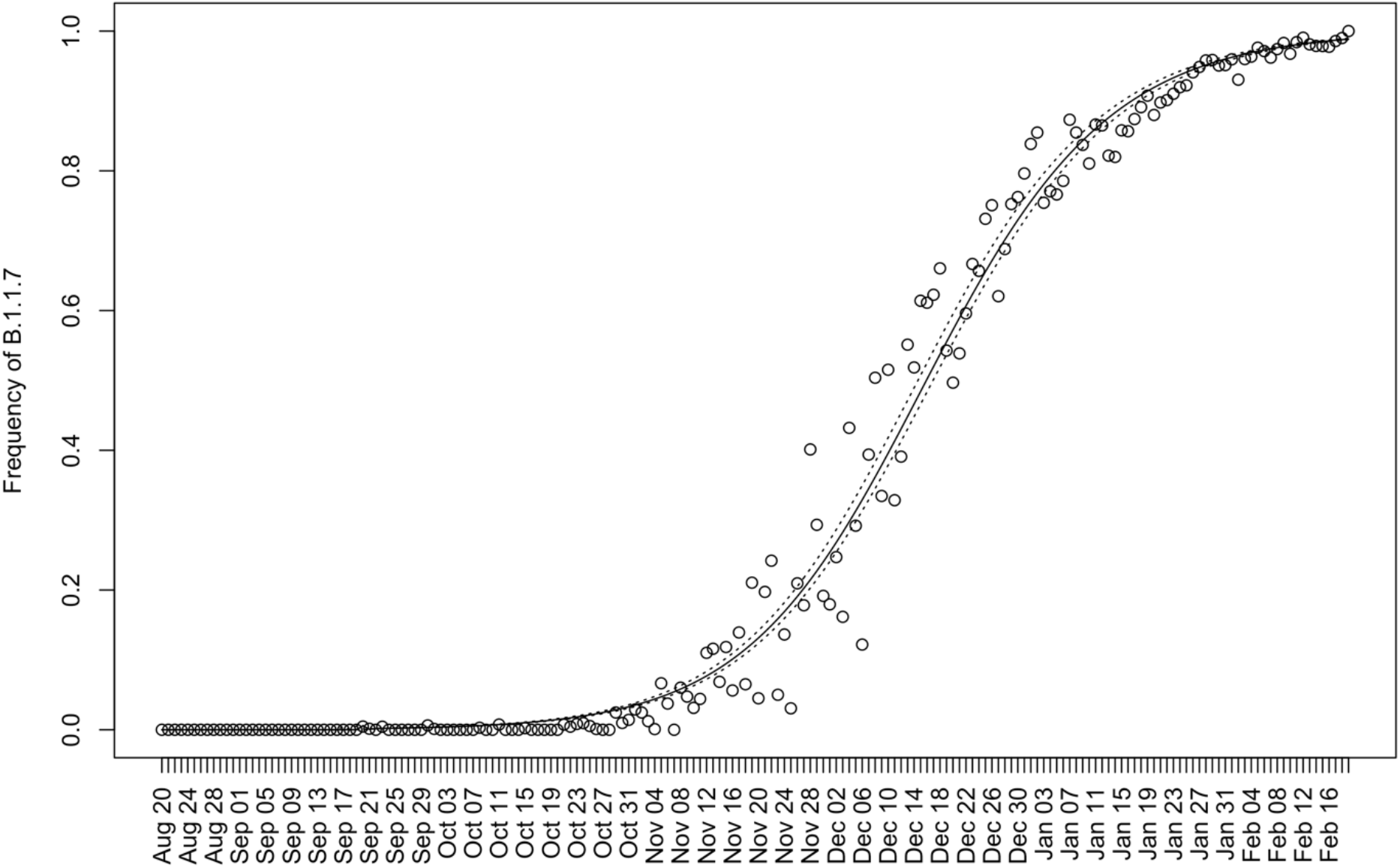
Time course of the frequency of B.1.1.7 strains among all strains except B.1.1.7-like strains detected in England from September 1, 2020 to February 19, 2021. White circles indicate the frequency of B.1.1.7 strains among B.1.1.7 and non-B.1.1.7 strains. The nucleotide sequences were retrieved from GISAID on March 1, 2021. Solid line indicates the time course of frequency of B.1.1.7 strains calculated using parameters estimated from the data. Dashed lines indicate its lower and upper bounds of its 95% CI.

Figure 3 shows result of sensitivity analysis of selective advantage *s*. The maximum likelihood estimate of *s* was affected by the log mean in a linear manner (Figure 3A). The minimum and maximum values of *s* on the ovals in Panel A and Panel B in Figure 3 were 0.266 and 0.459, respectively. From this result, we can conclude that the selective advantage *s* of B.1.1.7 strain over previous strains in England was between 0.266 and 0.459.

**Figure 3.**
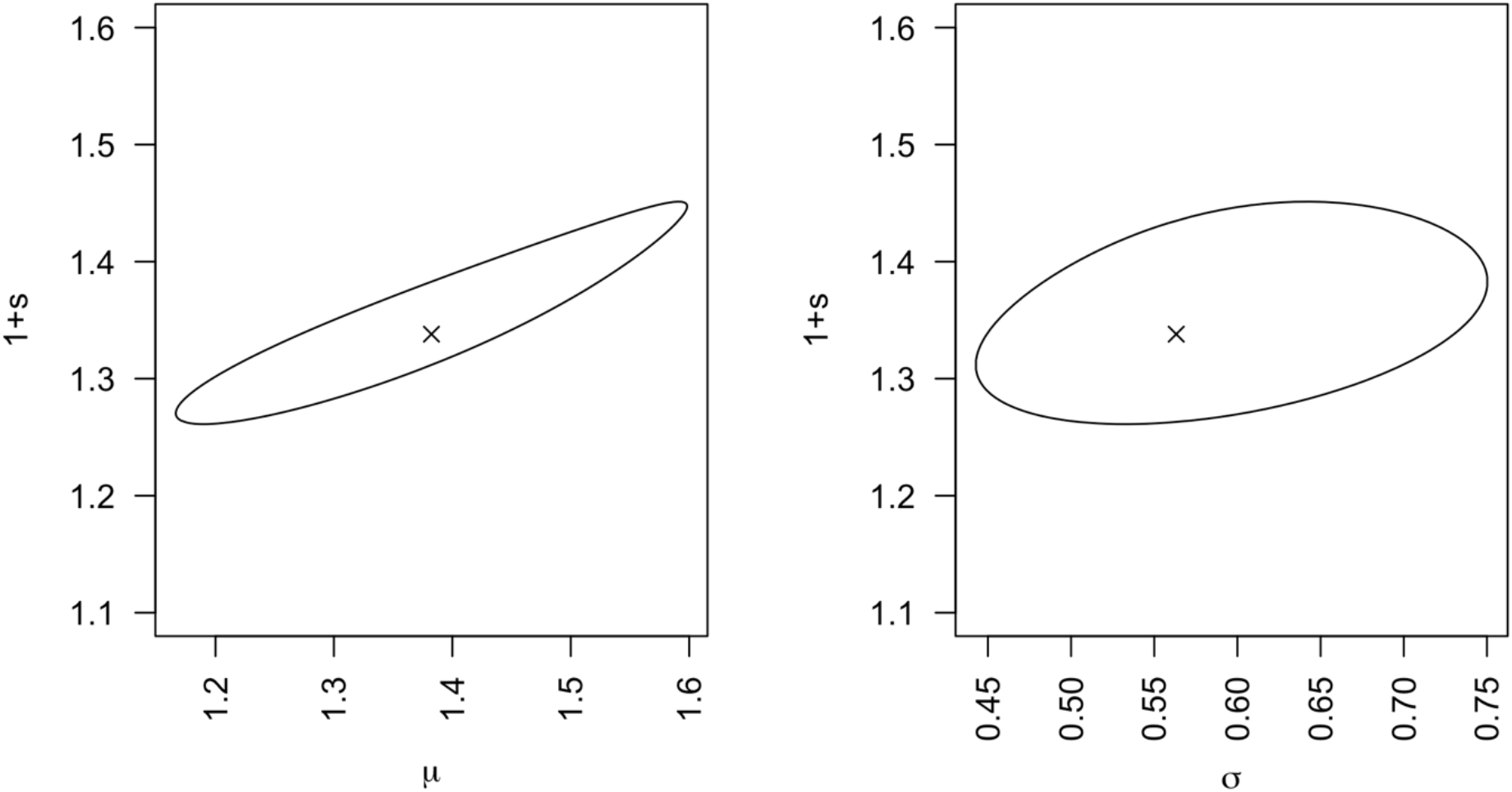
Effects of log mean (A) and log standard deviation (B) of serial interval distribution on the estimate of selective advantage, *s*. The cross marks in both panels represent the maximum likelihood estimate of *s* when the serial interval distribution was assumed to be a lognormal distribution with a log mean of 1.38 and log standard deviation of 0.563, which were estimated by Nishiura et al. (Nishiura et al., 2020). Areas inside oval in Panels A and B represent the range of maximum likelihood estimate of *s* obtained by assuming mean and standard deviation within the 95% confidence area shown in the Supplementary Figure 1.

Figure 4 shows the temporal change in the average **1** + ***s*** in the viral population circulating in England from September 1, 2020 to February 19, 2021. The value of **1** + ***s*** stayed around 1 until the end of October, 2020. After November, 2020, the average **1** + ***s*** in the viral population kept increasing due to the of increasing frequency of the B.1.1.7 strain. The increase leveled off around the end of January 2021, when the preexisting strain went extinct.

**Figure 4.**
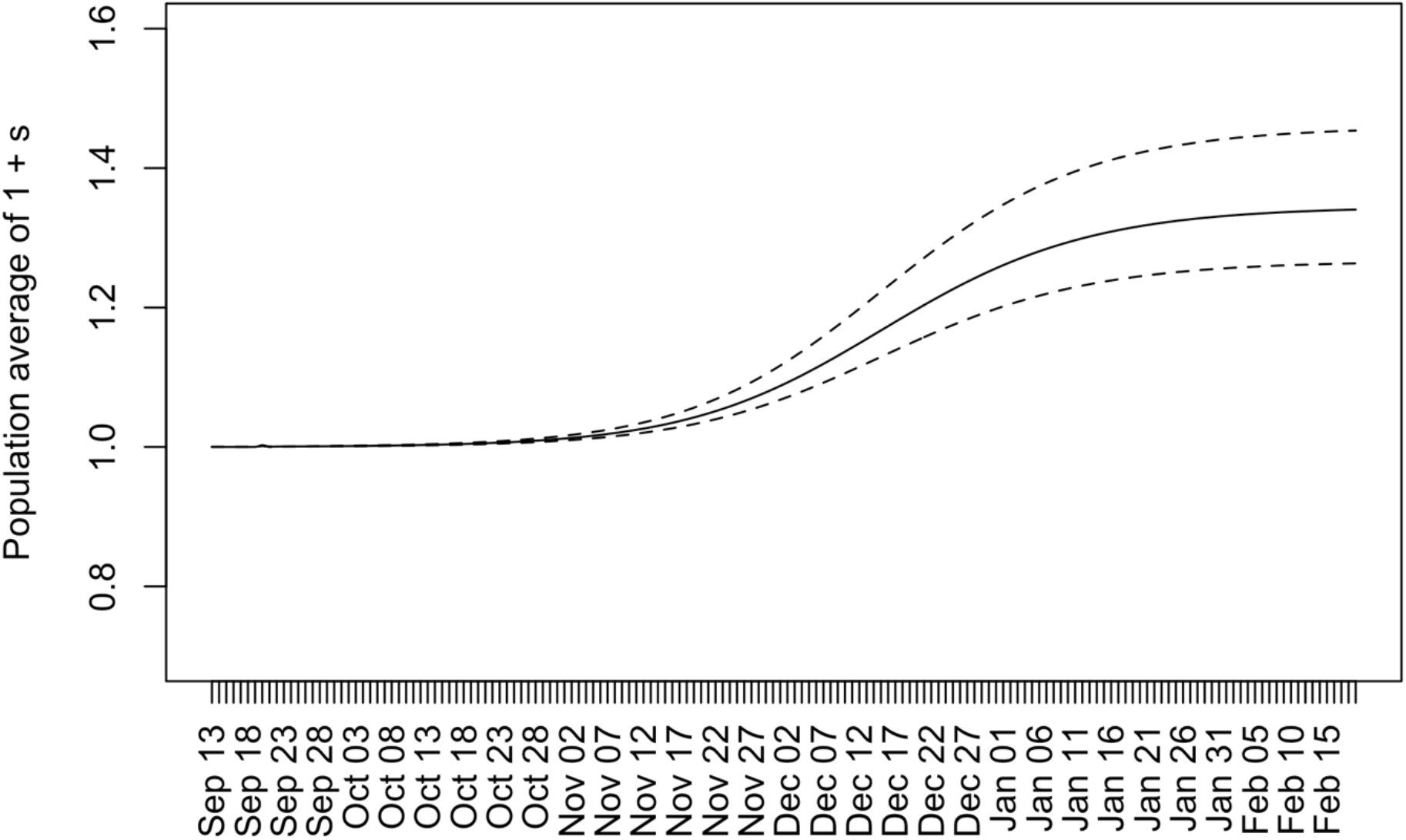
The temporal change in the average of 1 + *s* in the viral population circulating in England. The solid line indicates the population average of 1 + *s* when *s* =0.344 which was calculated using maximum likelihood estimation of the lognormal serial interval distribution. The dashed lines indicate the population average of 1 + *s* when *s* = 0. 266 (lower) and *s* = 0. 459 (upper), which are calculated via a sensitivity analysis.

## Discussion

In this paper, the selective advantage of the B.1.1.7 strain over non-B.1.1.7 strains in England was estimated to be 0.344 with a 95% CI from 0.343 to 0.346, assuming that the serial intervals followed the lognormal distribution with a log mean of 1.38 and log standard deviation of 0.56. The date of emergence of B.1.1.7 strains in England was estimated to be September 20, 2020 with its 95% confidence interval from September 17, 2020 to September 20, 2020. The initial frequency of B.1.1.7 among all sequences except B.1.1.7-like strains at the time of emergence in England was estimated to be 0.00556 with a 95% confidence interval from 0.00534 to 0.00581. The sensitivity analyses showed that the estimate of selective advantage was affected by parameters of assumed lognormal serial interval distribution.

Accounting for the serial interval distribution, the instantaneous reproduction number of B.1.1.7 strain were estimated to be 26.6–45.9% higher than previous strains circulating in England.

Our analyses showed that the B.1.1.7 strain possesses 26.6–45.9% higher transmissibility compared to previously circulating strains in England. This result suggests that control measures should be strengthened by 26.6–45.9% when the B.1.1.7 is newly introduced in a country where viruses with similar transmissibility to the preexisting strain in England are predominant.

Our estimate is lower than some previously published estimates. For example, Volz et al. estimated a 50–100% increase in the reproduction number using PCR data from England (Volz et al. 2021). The reason for this discrepancy could be the difference in the assumed serial interval distributions. In this paper, we used the serial interval data published by Nishiura et al. (Nishiura et al. 2020). Other groups used different datasets, and there is some variation between these estimated values (Rai et al. 2021). Volz et al. assumes a generation time distribution with a mean of 6.4 days based on the results by Bi et al. (Bi et al. 2020). However, Ali et al. have reported that the serial interval estimated using data from China before January 22, 2020 was longer than estimates after January 22, 2020 (Ali et al. 2020). The serial interval estimated by Bi et al. contains data from before January 22, 2020 and there might be some possibility that the estimated serial interval does not reflect the current situation. This important limitation originates from the uncertainty surrounding serial interval distribution.

Our analysis assumes that samples are collected from a well-mixed population in England. However, the situation may vary from region to region in England. Several observed frequencies in Figure 2 were located outside the 95% confidence interval. The reason for this could be that samples were collected from different locations in England and regional difference in the viral population may be the cause of the fluctuation of observed frequencies.

Our estimation method is based on the principle that the expected frequency of a mutant strain among all strains can be determined from those in the previous generation using the serial interval distribution of infections. The method assumes that the selective advantage of a mutant strain over previously circulating strains is constant over time, which is based on Maynard Smith’s model of allele frequencies in adaptive evolution. In line with Fraser’s method for estimating the instantaneous reproduction number, our method allows reproduction numbers of strains to change during the target period of analysis. Thus, the proposed method removes the assumption that the reproduction number is constant over time, which is assumed in previous studies. Our method can estimate the selective advantage of viruses in a genotype over the other genotype without estimating the reproduction numbers of viruses of each genotype. Thus, the method can be applicable for the analysis on the selection of new variants even when strong control measures such as lockdown were introduced during the target period of analysis. We think this is the largest contribution of this paper to the field of molecular evolution, population genetics, and infectious disease epidemiology.

As of June 9, 2021, the B.1.1.7 strain has been detected in 135 countries (Rambaut et al. 2020a). Estimation of the selective advantage of the B.1.1.7 strains over previously circulating strains in other countries is ongoing. Variant strains originating from Brazil, South Africa, and India also show higher transmissibility compared to previously circulating strains (World Health Organization 2021). There is an urgent need to estimate the selective advantage of these strains. We hope that the methodology developed in this paper proves useful for countries in the world to establish control measures against highly transmissible variants strains.

## Data Availability

Supplementary Table S1 contains the dataset used in the analysis.

## Acknowledgement

We gratefully acknowledge the laboratories responsible for obtaining the specimens and the laboratories where genetic sequence data were generated and shared via the GISAID Initiative, on which this research is based. This work was supported by the Japan Agency for Medical Research and Development (grant numbers JP20fk0108535). The work was also supported by the Grant-in-Aid (grant number 21H03490) and by the World-leading Innovative and Smart Education Program (1801) both from the Ministry of Education, Culture, Sports, Science, and Technology, Japan. The funders had no role in the study design, data collection and analysis, decision to publish, or preparation of the manuscript.

## Supplementary Materials

**Supplementary Figure 1.**
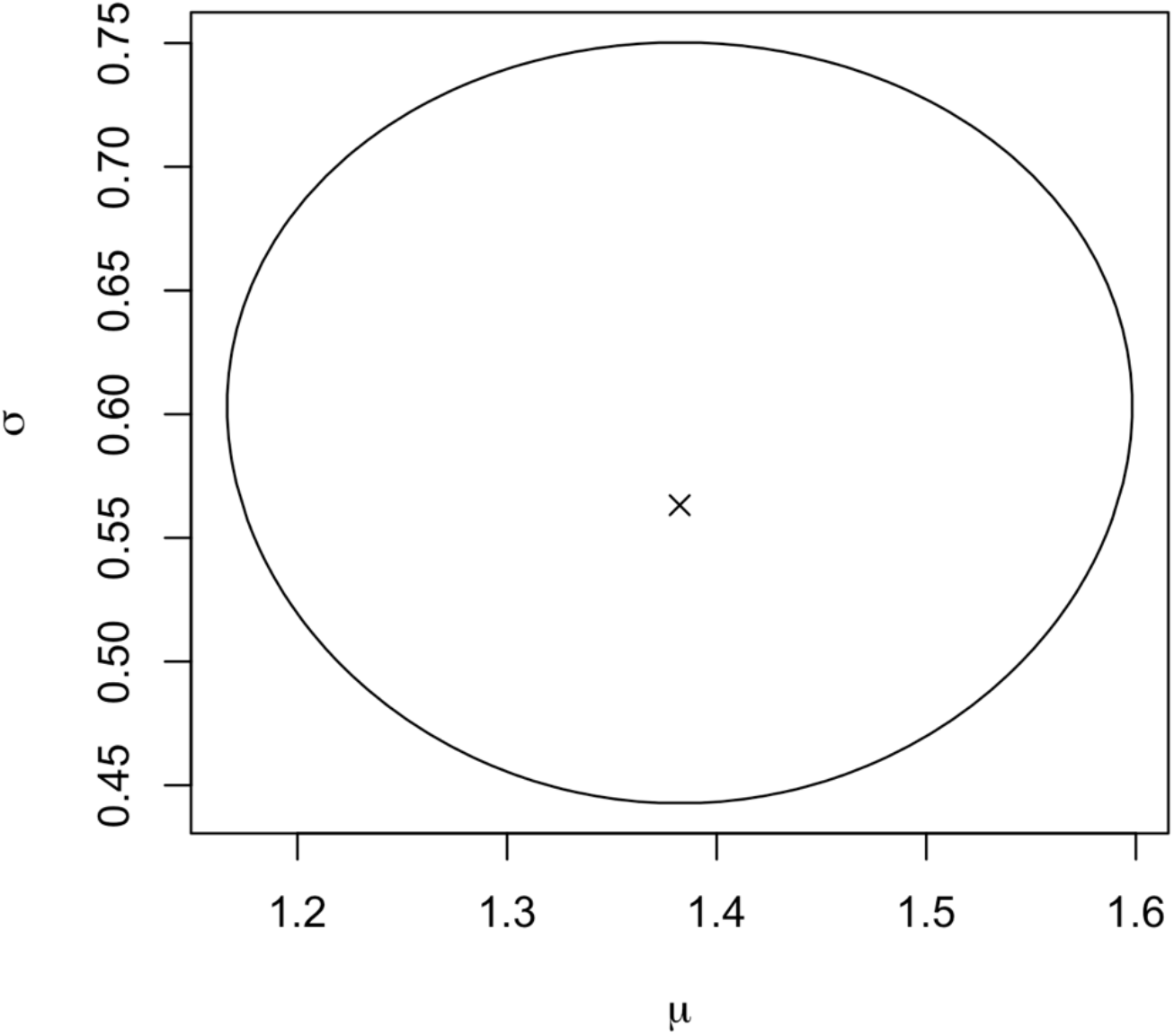
The 95% confidence area of log mean *μ* and log standard deviation *σ* of the lognormal distribution estimated using data obtained by Nishiura et.al (Nishiura et al. 2020). The cross mark represents the point of maximum likelihood estimates of *μ* and *σ*. A point inside the oval falls within the 95% confidence intervals.

